# Clinical Validation of a Proteomic Biomarker Threshold for Increased Risk of Spontaneous Preterm Birth

**DOI:** 10.1101/2021.01.23.21249902

**Authors:** Julja Burchard, Ashoka D. Polpitiya, Angela C. Fox, Todd Randolph, Tracey C. Fleischer, Max Dufford, Thomas J. Garite, Greg C. Critchfield, J. Jay Boniface, Paul E. Kearney

**Affiliations:** Sera Prognostics, Inc.; Sera Prognostics, Inc., independent consultant; Data Incites, Inc.

## Abstract

Preterm births are prevalent and a leading cause of neonatal death in the United States. Despite the availability of effective interventions, to date there is not a robust and widely applicable test to identify pregnancies at high risk for spontaneous preterm birth (sPTB). Previously, a sPTB predictor based on the ratio of two proteins, IBP4/SHBG, was validated as an accurate predictor of sPTB in the observational study Proteomic Assessment of Preterm Risk (PAPR). Here it is demonstrated that the same predictor threshold associated with 2-fold increased risk of sPTB, namely −1.4, is also statistically significant for predicting elevated risk of sPTB in the observational study Multicenter Assessment of a Spontaneous Preterm Birth Risk Predictor (TreeToP).

## Introduction

Preterm birth (PTB) occurs in 10% of all births and is the second leading cause of neonatal death in the United States.^1,2^ The effective application of interventions, such as progesterone, to help prevent PTB and its sequelae requires diagnostic tools to identify pregnancies at risk. Clinical markers for assessing the risk of PTB are in general poorly predictive and are present in only a small minority of pregnancies. A history of previous PTB is a traditional predictor of recurrent PTB but is seen in only approximately 4% of all PTBs.^3,4^ Similarly, a short cervical length measured by transvaginal ultrasound is a predictor of PTB, but accounts for only an additional 2% of all PTBs.^5,6^ Of particular interest are diagnostic tools that predict spontaneous PTB (sPTB) which accounts for approximately 60% of all PTBs.^7^ Saade and colleagues described and clinically validated a novel serum proteomic spontaneous PTB predictor based on the ratio of insulin-like growth factor-binding protein 4 (IBP4, gene symbol IGFBP4) to sex hormone-binding globulin (SHBG) using samples from the Proteomic Assessment of Preterm Risk (PAPR) study (NCT01371019).^8^ Additionally, the protein ratio IBP4/SHBG test has been analytically validated.^9^ We refer to this as the “protein ratio” or “test” throughout this document.

In accordance with the National Academy of Medicine’s guidelines^10^ for the rigorous development of molecular tests, we clinically validate that when the protein ratio IBP4/SHBG is above a specific threshold there is a statistically significant higher risk of sPTB. It is desirable for a diagnostic test to risk stratify subjects at a specified threshold so that physicians can interpret and act upon test results. For example, in the context of sPTB, to apply preventative measures to reduce the risk that a pregnancy will result in sPTB when a test score is above the threshold.

Specifically, in this work we will demonstrate that the protein ratio IBP4/SHBG, which demonstrated a statistically elevated risk of sPTB at threshold −1.4 in the PAPR study, and also demonstrated a statistically elevated risk of sPTB at threshold −1.4 in the Multicenter Assessment of a Spontaneous Preterm Birth Risk Predictor (TreeToP) study (NCT02787213).^11^ This demonstrates the reproducibility of the protein ratio threshold in two large observational studies for predicting elevated risk of sPTB.

## Methods

### PAPR and TreeToP Subpopulation Selection

Subpopulations of PAPR and TreeToP were selected to conduct the analysis as described below. We note that all subjects are within the intended use population of the protein ratio test.

The subpopulation of PAPR used in this analysis consists of those subjects (n=549) who did not receive progesterone after 14 weeks gestation, underwent sample collection in the validated^8^ gestational age window (19^1/7^ − 20^6/7^ weeks) and gave consent for future research use of their deidentified samples and data.

The subpopulation of TreeToP used in this analysis consists of a randomly selected subset of approximately 30% of all subjects (n=847) who underwent sample collection in the validated gestational age window (19^1/7^ − 20^6/7^ weeks). These subjects were randomly selected by a third-party statistician for an interim analysis and reflect the clinical and demographic factors of the overall study. We note that that the non-selected subjects in the study remain blinded for future studies.

In the comparison of PAPR to TreeToP the published index scoring system, “0 to 4 scale with NICU”^6^ was adapted to measure neonatal composite morbidity and mortality (hNMI). For each neonate, the index increases from 1 to 3 for each additional diagnosis of respiratory distress syndrome, bronchopulmonary dysplasia, intraventricular hemorrhage grade III or IV, all stages of necrotizing enterocolitis, periventricular leukomalacia or proven severe sepsis. The published scale uses NICU stays to determine index scores if the length of stay gives a higher score than concomitant diagnoses: 1-4 days give a score of 1, 5-20 days a score of 2 and >20 days a score of 3. For this study, as PAPR did not record NICU stay the published scale was adapted to utilize hospital stay in place of NICU length of stay. Perinatal mortality (intrauterine fetal demise or neonatal mortality) is scored as 4. Data collection through 28 days of life allowed for confirmation of all conditions contributing to hNMI.

### Sample Analysis

All subject samples from PAPR and TreeToP were analyzed in a certified lab according to standard operating protocol using a methodology previously validated and documented.^9^ Importantly, the TreeToP samples were analyzed prospectively, as they were collected, as they would be in actual clinical use.

### Analysis Methodology

The protein ratio IBP4/SHBG threshold of −1.4 was pre-identified as an important clinical decision point as it corresponds to a 2-fold increase in risk of sPTB in the PAPR subjects. Specifically, it was demonstrated by Kaplan-Meier analysis that threshold −1.367 had statistical significance (p-value 0.0004) in a nested case-control cohort of PAPR.^8^ Re-analysis of threshold significance in a larger PAPR cohort is performed in this study.

The objective is to demonstrate that at the same protein ratio threshold (−1.367) there is also a statistically significant increase in sPTB risk in TreeToP, thereby demonstrating in an independent cohort of intended use subjects, that this protein ratio threshold is reproducibly significant for elevated sPTB risk. The results may further help determine the potential clinical utility of the biomarker in routine clinical practice.

As the PAPR and TreeToP sets of subjects are finite in size, the distribution of protein ratio thresholds will not be exactly the same. For this reason, the specific hypothesis that is tested on the TreeToP subjects is that there is a protein ratio threshold, in the range of −1.367 +/-0.114, that is statistically significant for increased risk of sPTB. It is also noted that in the previously reported analytical validation, the technology platform for measuring the protein ratio has a precision with standard deviation of 0.114.^9^ Statistical significance will be assessed as one-sided p-values from regression with BMI as a covariate, indicated by reported influence of BMI stratification in prediction of sPTB.^8^ The same test of significance is used in PAPR and TreeToP to avoid bias.

## Results

The PAPR and TreeToP subpopulations are described and contrasted in Table 1.

**Table 1.** A comparison of the PAPR and TreeToP populations analyzed on various demographic categories. P-value and the standardized mean difference (smd) are provided for statistical comparisons.

| Variable | PAPR | TreeToP | p-value | smd |
| --- | --- | --- | --- | --- |
| Maternal Education n (%) |  |  |  |  |
| Unknown | 2 (0.36) | 3 (0.35) | 0.000 | 0.404 |
| No grad | 144 (26.23) | 128 (15.11) |  |  |
| HS/GED | 278 (50.64) | 381 (44.98) |  |  |
| BA/S+ | 125 (22.77) | 335 (39.55) |  |  |
| Race n (%) |  |  |  |  |
| Black or African American | 89 (16.21) | 173 (20.43) | 0.000 | 0.335 |
| Other | 52 (9.47) | 165 (19.48) |  |  |
| White | 408 (74.32) | 509 (60.09) |  |  |
| Ethnicity n (%) |  |  |  |  |
| Hispanic or Latino | 192 (34.97) | 335 (39.55) | 0.095 | 0.119 |
| Non-Hispanic or Latino | 357 (65.03) | 510 (60.21) |  |  |
| Unknown | 0 (0.00) | 2 (0.24) |  |  |
| Gravida n (%) |  |  |  |  |
| Multigravida | 389 (70.86) | 570 (67.30) | 0.174 | 0.077 |
| Primigravida | 160 (29.14) | 277 (32.70) |  |  |
| Prior Full-term Birth n (%) |  |  |  |  |
| First Pregnancy | 160 (29.14) | 277 (32.70) | 0.289 | 0.086 |
| None | 76 (13.84) | 101 (11.92) |  |  |
| One or more | 313 (57.01) | 469 (55.37) |  |  |
| Prior sPTB n (%) |  |  |  |  |
| First Pregnancy | 160 (29.14) | 277 (32.70) | 0.000 | 0.351 |
| None | 324 (59.02) | 546 (64.46) |  |  |
| One or more | 65 (11.84) | 24 (2.83) |  |  |
| Delivery n (%) |  |  |  |  |
| miPTB | 29 (5.28) | 32 (3.78) | 0.027 | 0.145 |
| sPTB | 37 (6.74) | 34 (4.01) |  |  |
| Term | 483 (87.98) | 781 (92.21) |  |  |
| Fetal Gender n (%) |  |  |  |  |
| Ambiguous | 0 (0.00) | 1 (0.12) | 0.771 | 0.057 |
| Female | 282 (51.37) | 422 (49.82) |  |  |
| Male | 267 (48.63) | 424 (50.06) |  |  |
| Neonatal morbidity and mortality index n (%) |  |  |  |  |
| 1 | 482 (87.80) | 767 (90.55) | 0.083 | 0.138 |
| 2 | 50 (9.11) | 55 (6.49) |  |  |
| 3 | 10 (1.82) | 21 (2.48) |  |  |
| 4 | 7 (1.28) | 4 (0.47) |  |  |

In PAPR and TreeToP, a protein ratio threshold of −1.375 was statistically significant for increased sPTB (both at p-value 0.041) which is within the pre-specified range of −1.367 +/-.114.

To test the reproducibility of stratification demonstrated in clinical validation^8^, Kaplan-Meier curves for gestational age at birth in PAPR and TreeToP were generated at this threshold (see Figure 1 and 2). Stratification of gestational age at birth is significant by log-rank test with truncation at 40 weeks in both studies at this threshold (PAPR: p-value 5.2e-6; TREETOP: p-value 1.6e-3).

**Figure 1.**
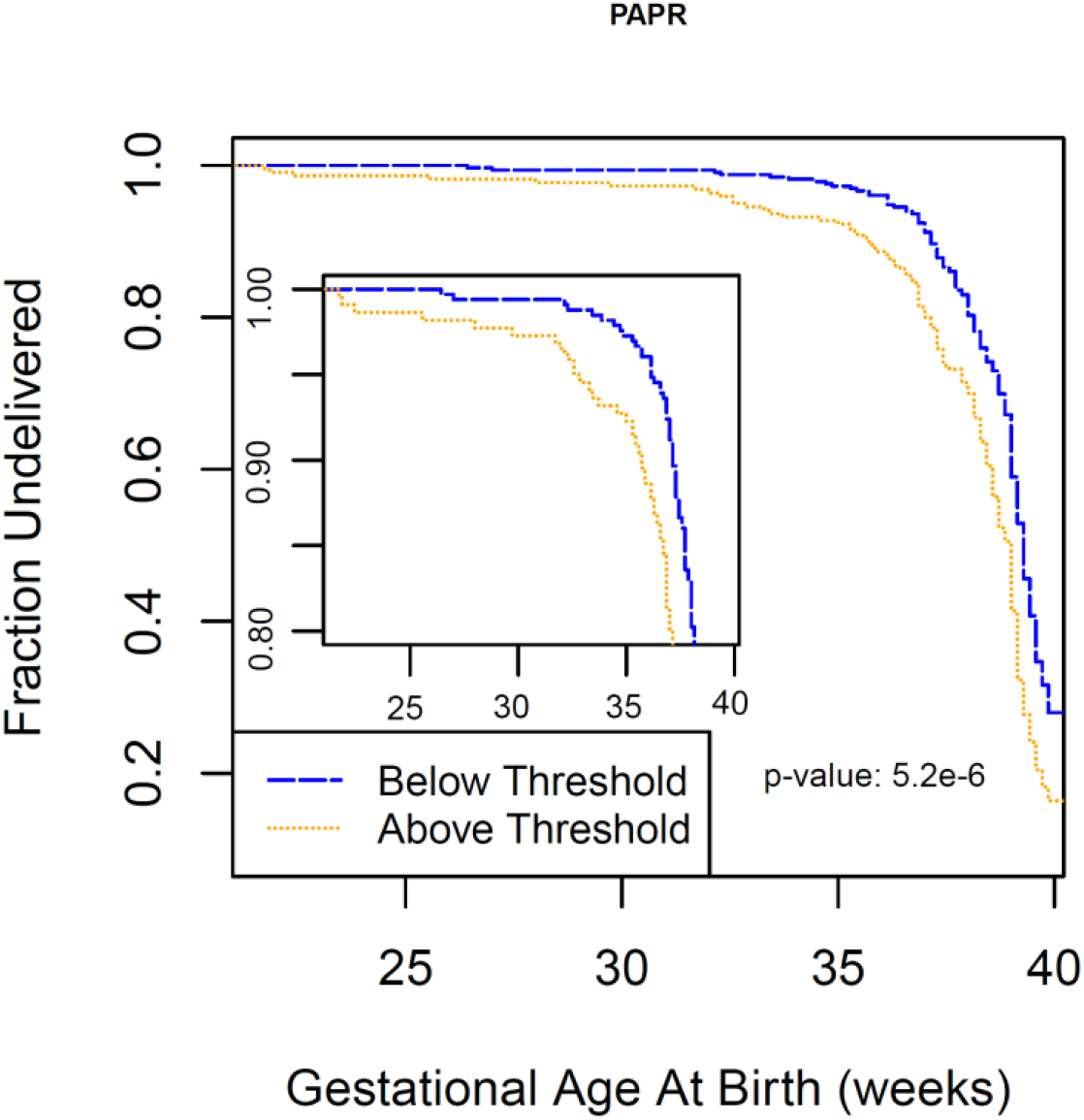
Kaplan-Meier curve for the PAPR study for all subjects up to 40 weeks of gestation and truncated at the earliest 20% of deliveries (inset) stratified by the protein ratio −1.375.

**Figure 2.**
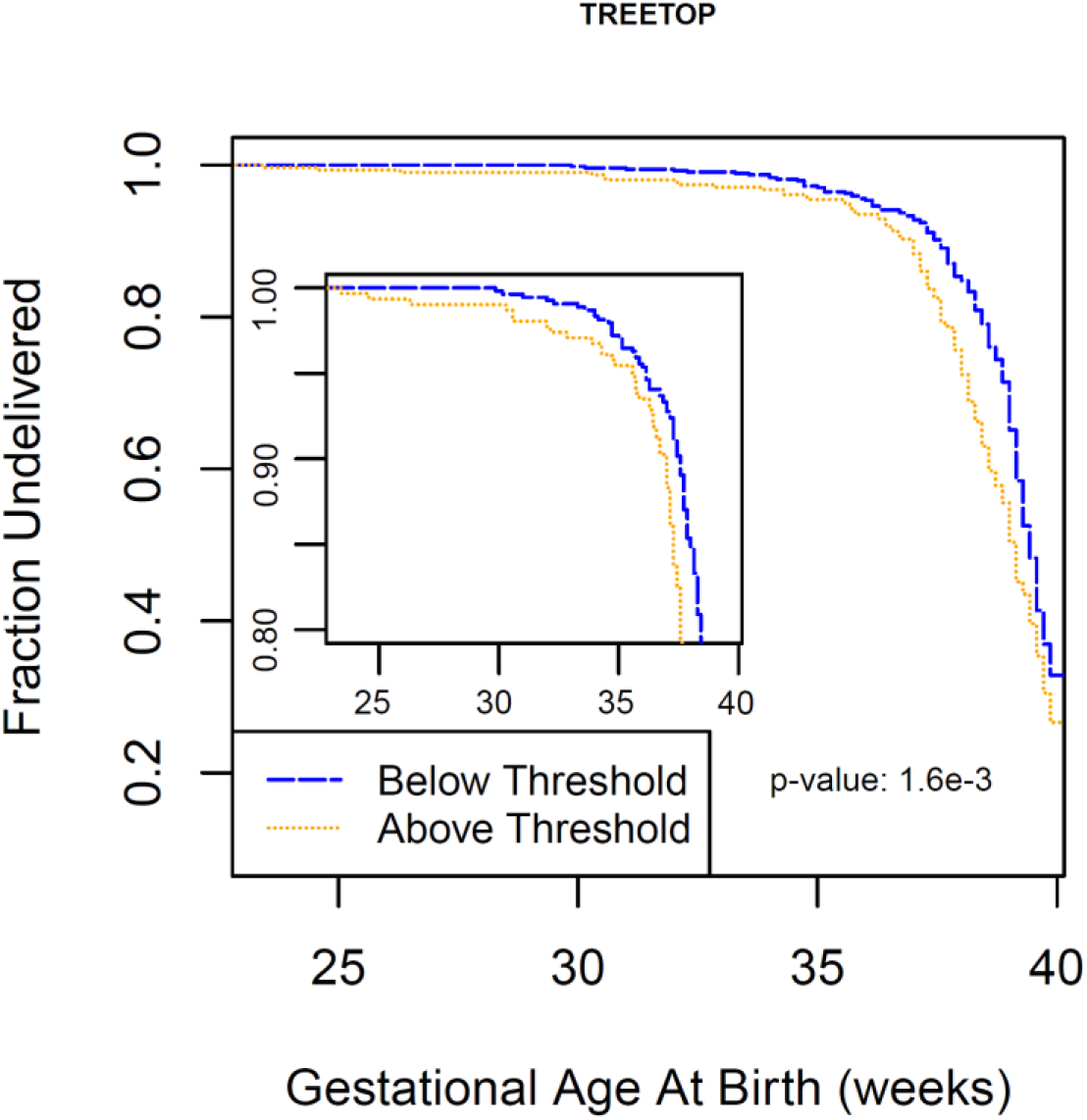
Kaplan-Meier curve for the TREETOP study for all subjects up to 40 weeks of gestation and truncated at the earliest 20% of deliveries (inset) stratified by the protein ratio −1.375.

## Discussion

The primary outcome of this research was to demonstrate that the protein ratio threshold corresponding to a two-fold increased risk of sPTB in the PAPR study is also statistically significant in the TreeToP study for increased risk of sPTB. That is, the IBP4/SHBG protein ratio threshold of −1.4 has been clinically validated for elevated sPTB.

Although both the PAPR and TreeToP subpopulations analyzed are intended use populations for the validated protein ratio test, they are notably different on several demographic parameters (education, race, prior sPTB, etc.). Additionally, the PAPR and TreeToP studies enrolled subjects at 11 and 18 sites, respectively. Despite these demographic differences and diversity in site enrolment, the same protein ratio threshold identifies pregnancies of increased risk of sPTB. This is strong evidence of the robust reproducibility and generalizability of the threshold.

## Data Availability

All data generated for this manuscript is available by contacting

## Acknowledgements

We thank the Principal Investigators of the PAPR (NCT01371019) and TreeToP (NCT02787213) clinical trials for their support. We are indebted to the feedback received by Drs. George R. Saade and Glenn R. Markenson.

